# Comparative Efficacy and Safety of De-escalation, Abbreviation, and Standard Potent P2Y_12_ Inhibitor–Based Dual Antiplatelet Therapy Strategies After Acute Coronary Syndrome: A Network Meta-Analysis

**DOI:** 10.1101/2025.11.16.25339537

**Authors:** Yee-Jen Wu, Hung-Ju Lin, Yi-Chen Chou, Yen-Hung Lin, Chi-Sheng Hung

**Author notes:** **Corresponding author:** Chi-Sheng Hung, Division of Cardiology, Department of Internal Medicine and Cardiovascular Center, National Taiwan University Hospital, Taipei, Taiwan Address: 100, 7 Chung Shan S. Rd., Taipei, Taiwan.

## Abstract

**Background:** The efficacy and safety of de-escalation from potent P2Y_12_ inhibitor-based dual antiplatelet therapy (DAPT) to clopidogrel-based DAPT, or abbreviation to potent P2Y_12_ inhibitor monotherapy, compared with standard 12-month DAPT, remain unclear after acute coronary syndrome (ACS).

**Methods:** Frequentist and Bayesian network meta-analyses of randomized controlled trials were performed to compare three guideline-endorsed strategies: (1) short-term potent P2Y_12_ inhibitor–based DAPT de-escalation to clopidogrel-based DAPT, (2) short-term potent P2Y_12_ inhibitor–based DAPT and abbreviated DAPT followed by potent P2Y_12_ inhibitor monotherapy, and (3) standard 12-month potent P2Y_12_ inhibitor–based DAPT. The primary efficacy endpoint was major adverse cardiovascular events (MACEs). The key secondary endpoint was net adverse clinical events (NACEs). The primary and secondary safety endpoints were major bleeding and clinically relevant bleeding, respectively.

**Results:** Seven randomized controlled trials involving 20,673 patients were included. Both the de-escalation and abbreviation strategies significantly reduced major bleeding (RR 0.43, 95%CI 0.25-0.74, p=0.002; and RR 0.43, 95%CI 0.33-0.58, p<0.001, respectively) and NACEs (RR 0.54, 95%CI 0.41-0.70, p<0.001; and RR 0.72, 95%CI 0.61-0.84, p<0.001, respectively) without increasing MACEs or other ischemic outcomes. Indirect comparison indicated that de-escalation vs abbreviation strategies demonstrated comparable MACE and bleeding outcomes.

**Conclusions:** Following ACS, de-escalation from potent P2Y_12_ inhibitor-based DAPT to clopidogrel-based DAPT or abbreviation to potent P2Y_12_ inhibitor monotherapy can reduce bleeding risk without compromising ischemic protection. While there were no significant differences in MACEs or NACEs between the two simplified strategies, further research is warranted to identify the most appropriate individualized strategy.

## Introduction

Acute coronary syndrome (ACS) remains a major cause of morbidity and mortality worldwide. Dual antiplatelet therapy (DAPT) with aspirin plus a P2Y_12_ inhibitor is the standard of care after percutaneous coronary intervention (PCI). Current guidelines recommend 12 months of DAPT, with a preference toward potent P2Y_12_ inhibitors.^1,2^ However, prolonged DAPT use increases the risk of bleeding, which has prognostic implications similar to those of ischemic complications.^3^ This tradeoff has driven the evaluation of alternative antiplatelet strategies after ACS.

Recent randomized controlled trials (RCTs) have indicated that abbreviating DAPT to ≤3 months followed by P2Y_12_ inhibitor monotherapy may provide comparable ischemic protection and significantly reduced bleeding events, with ticagrelor monotherapy supported by the most consistent evidence.^4–8^ Alternatively, de-escalation from a potent P2Y_12_ inhibitor to clopidogrel has also demonstrated favorable safety and efficacy outcomes.^9,10^ Several meta-analyses have attempted to compare the effects of de-escalation and abbreviation strategies after ACS.^11–16^

However, the interpretations of some of these analyses have been limited by the inclusion of both potent P2Y_12_ inhibitors and clopidogrel or even aspirin alone in the same treatment arm.^11,13–16^ Therefore, whether abbreviation to potent P2Y_12_ inhibitor monotherapy or de-escalation to aspirin plus clopidogrel–based DAPT yields the most favorable net clinical outcomes after a short initial course of potent P2Y_12_ inhibitor–based DAPT remains uncertain.

To address this knowledge gap, we conducted a systematic review and network meta-analysis to clarify the comparative effectiveness and safety of the aforementioned strategies. Our findings may have direct implications for tailoring antiplatelet therapy in contemporary ACS care.

## Methods

We conducted this systematic review and network meta-analysis according to the Preferred Reporting Items for Systematic Reviews and Meta-Analyses (PRISMA) statement. In line with PRISMA recommendations, the research question was structured using the Population, Intervention, Comparison, and Outcomes (PICO) framework (Table 1). This study was prospectively registered at PROSPERO (CRD420251144892). Institutional ethics committee approval was waived because of the nature of the study.

**Table 1.** PICO Framework.

|  |  |
| --- | --- |
| **Population** | Patients with ACS undergoing PCI |
| **Intervention** | De-escalation strategy: Potent P2Y<sub>12</sub> inhibitor–based DAPT for 1 to 3 months, followed by clopidogrel-based DAPT until 12 months after ACS<br>Abbreviation strategy: Potent P2Y<sub>12</sub> inhibitor–based DAPT within 3 months, followed by potent P2Y<sub>12</sub> inhibitor monotherapy until 12 months after ACS |
| **Comparator** | Standard strategy: Potent P2Y<sub>12</sub> inhibitor–based DAPT for 12 months after ACS |
| **Outcome** | Efficacy outcomes:<br>• All-cause mortality<br>• Cardiovascular mortality<br>• MI<br>• Stent thrombosis<br>• Stroke<br>• MACEs<br>2) Safety outcomes:<br>• Major bleeding<br>• Clinically relevant bleeding<br>3) NACEs |

### Data Source and Search Strategy

PubMed, EMBASE, Web of Science, and Cochrane CENTRAL were searched for relevant studies, supplemented by a manual screening of reference lists from previous studies and reviews. Only full-text articles published in English until September 20, 2025, were included. The detailed search strategy is outlined in the Supplementary Material. In addition, the reference lists of relevant articles were manually screened to identify any additional eligible studies.

### Study Selection

This systematic review and network meta-analysis included RCTs comparing strategies for de-escalation from short-term DAPT to clopidogrel plus aspirin, abbreviated DAPT followed by potent P2Y_12_ inhibitor monotherapy, and standard DAPT after PCI in patients with ACS. The inclusion criteria were as follows: having an RCT design, involving patients presenting with ACS, comparing between potent P2Y_12_ inhibitor monotherapy after short-term (≤3 months) DAPT as the experimental group and potent P2Y_12_ inhibitor–based standard (12 months) DAPT as the control group or comparing between clopidogrel-based DAPT after short-term (≤3 months) DAPT as the experimental group and potent P2Y_12_ inhibitor–based standard (12 months) DAPT as the control group, and involving a follow-up duration of ≥12 months. The exclusion criteria were as follows: having a non-RCT design, not reporting the outcomes of patients with ACS, and involving patients with an indication for oral anticoagulants.

Potentially eligible studies were screened by 2 investigators (YJW and CSH). Any disagreements were discussed and resolved through consensus, followed by discussion with a third reviewer (HJL) when necessary.

### Data Extraction and Quality Assessment

Two investigators (YJW and CSH) independently extracted data from eligible studies. These data included publication year, study country, study acronym, study design, eligibility criteria, basic demographic and clinical characteristics, follow-up duration, efficacy outcomes, and safety outcomes. Any disagreements were discussed and resolved through consensus, followed by discussion with a third reviewer (HJL) when necessary. No ethical approval was required for this study. The methodological quality of the included RCTs was evaluated using the Cochrane Collaboration risk-of-bias tool for random sequence generation, allocation concealment, blinding, incomplete outcome data, selective outcome reporting, and other potential sources of bias.

### Endpoints

The primary efficacy endpoint was defined as major adverse cardiovascular events (MACEs), a composite of cardiovascular death, myocardial infarction (MI), or stroke. Individual MACE components were also separately analyzed. The key secondary efficacy endpoint was defined as net adverse clinical events (NACEs), a composite of ischemic events and bleeding events. The primary safety endpoint was identified as major bleeding, defined as Bleeding Academic Research Consortium (BARC) type 3 or 5 bleeding. When this information was unavailable, major bleeding was alternatively defined as BARC type 3 or 5 bleeding or as Thrombolysis in Myocardial Infarction (TIMI) major or minor bleeding, as appropriate. The secondary safety endpoint was identified as clinically relevant bleeding, defined as BARC type 2, 3, or 5 bleeding. When this information was unavailable, clinically relevant bleeding was alternatively defined as BARC type 2, 3, 4, or 5 bleeding or TIMI major or minor bleeding, as appropriate.

### Data Synthesis and Statistical Analysis

We compared 3 guideline-endorsed post-PCI antiplatelet strategies for patients with ACS (Figure S1):

1. De-escalation strategy (de-escalation to clopidogrel-based DAPT), including potent P2Y_12_ inhibitor–based DAPT for 1 to 3 months followed by clopidogrel-based DAPT to complete 12 months of treatment after ACS.
2. Abbreviation strategy (abbreviated DAPT followed by potent P2Y_12_ inhibitor monotherapy), including potent P2Y_12_ inhibitor–based DAPT within 3 months followed by potent P2Y_12_ inhibitor monotherapy to complete 12 months of treatment after ACS.
3. Standard strategy, including potent P2Y_12_ inhibitor–based DAPT for 12 months after ACS.

A network meta-analysis of study outcomes was conducted using a frequentist approach, with the 3 aforementioned strategies serving as network nodes. The results are reported as risk ratios (RRs) with corresponding 95% confidence intervals (CIs). Statistical heterogeneity was assessed through τ^2^ and I^2^ statistics. I^2^ values of <25%, 25%-50%, and >50% indicated low, moderate, and high heterogeneity, respectively. For each direct comparison, log RRs (or log hazard ratios) were pooled using a random-effects model.

Arm-based Bayesian network meta-analyses were conducted with a random-effects consistency model in R (gemtc and rjags packages). Four parallel Markov chain Monte Carlo chains were run with overdispersed initial values. After a burn-in phase of 50⍰000 iterations, an additional 200⍰000 iterations were retained for posterior estimation. Effects are reported as posterior median RRs with 95% credibility intervals (CrIs). Rank probabilities, rankograms, and surfaces under cumulative ranking curves were calculated to evaluate the relative ranking of each strategy.

Sensitivity analyses were conducted according to the duration of DAPT (ie, within 1 or 3 months in the potent P2Y_12_ inhibitor monotherapy arm). Publication bias risk was estimated using funnel plots and Egger’s test. All analyses were conducted using R software version 4.5.0 (R Foundation for Statistical Computing, Vienna, Austria).

## Results

### Study Selection

Seven RCTs meeting the inclusion criteria were included in this network meta-analysis. Figure 1 depicts the geometry of the treatment network, which comprises 3 nodes. 7 RCTs evaluating the abbreviation strategy, 2 RCTs evaluating the de-escalation strategy, and 9 RCTs evaluating the standard strategy. No closed loop is observed in the network because of the lack of direct comparisons between the abbreviation and de-escalation strategies. Because the network structure comprises only star-shaped connections without closed loops, we could not perform local inconsistency assessments, such as node-splitting analyses.

**Figure 1.**
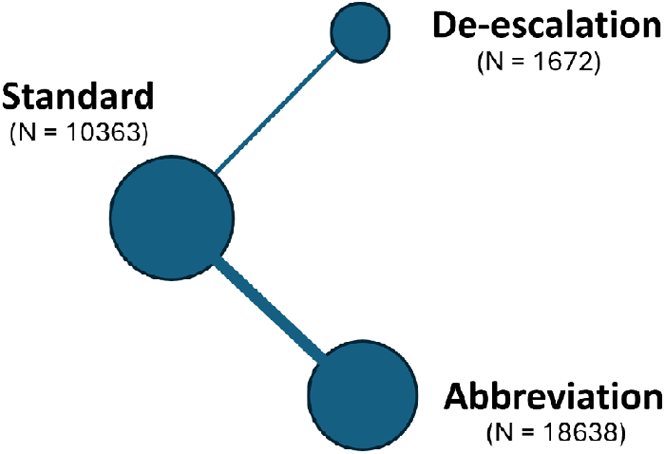
Geometry of the Treatment Network

### Study Description and Risk-of-Bias Assessment Results

Table 2 presents a summary of the characteristics of the included studies. All studies exclusively included patients with ACS. Randomization was performed within 3 months of potent P2Y_12_ inhibitor–based DAPT after the index ACS event date. The duration of follow-up was at least 12 month after the index ACS event date. Table S1 presents the baseline characteristics of each strategy. The ages of the included patients ranged from 58 to 64 years, with men representing 70% to 83% of the entire sample. Table S2 lists the definition of each outcome in the included RCTs.

**Table 2.**
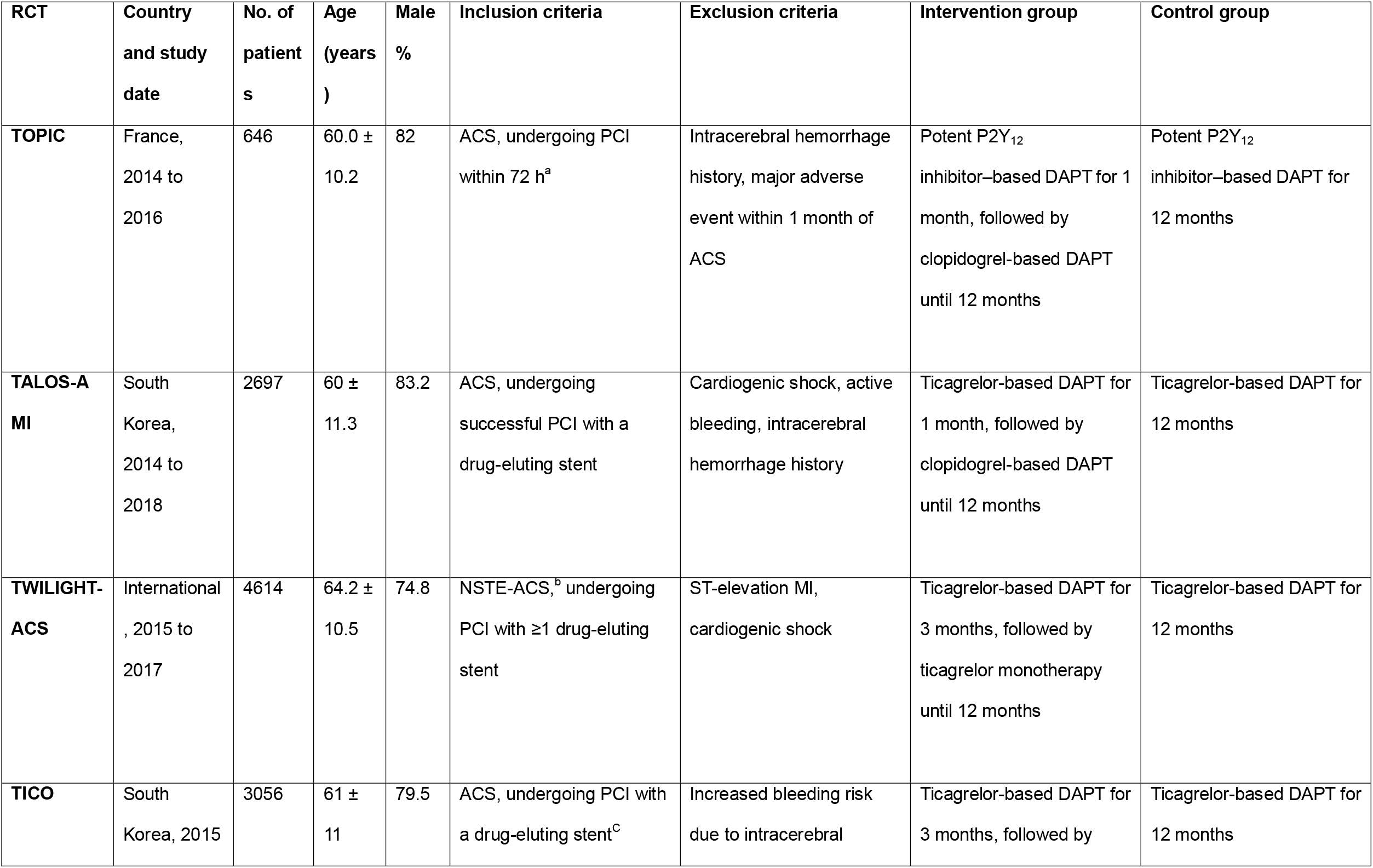
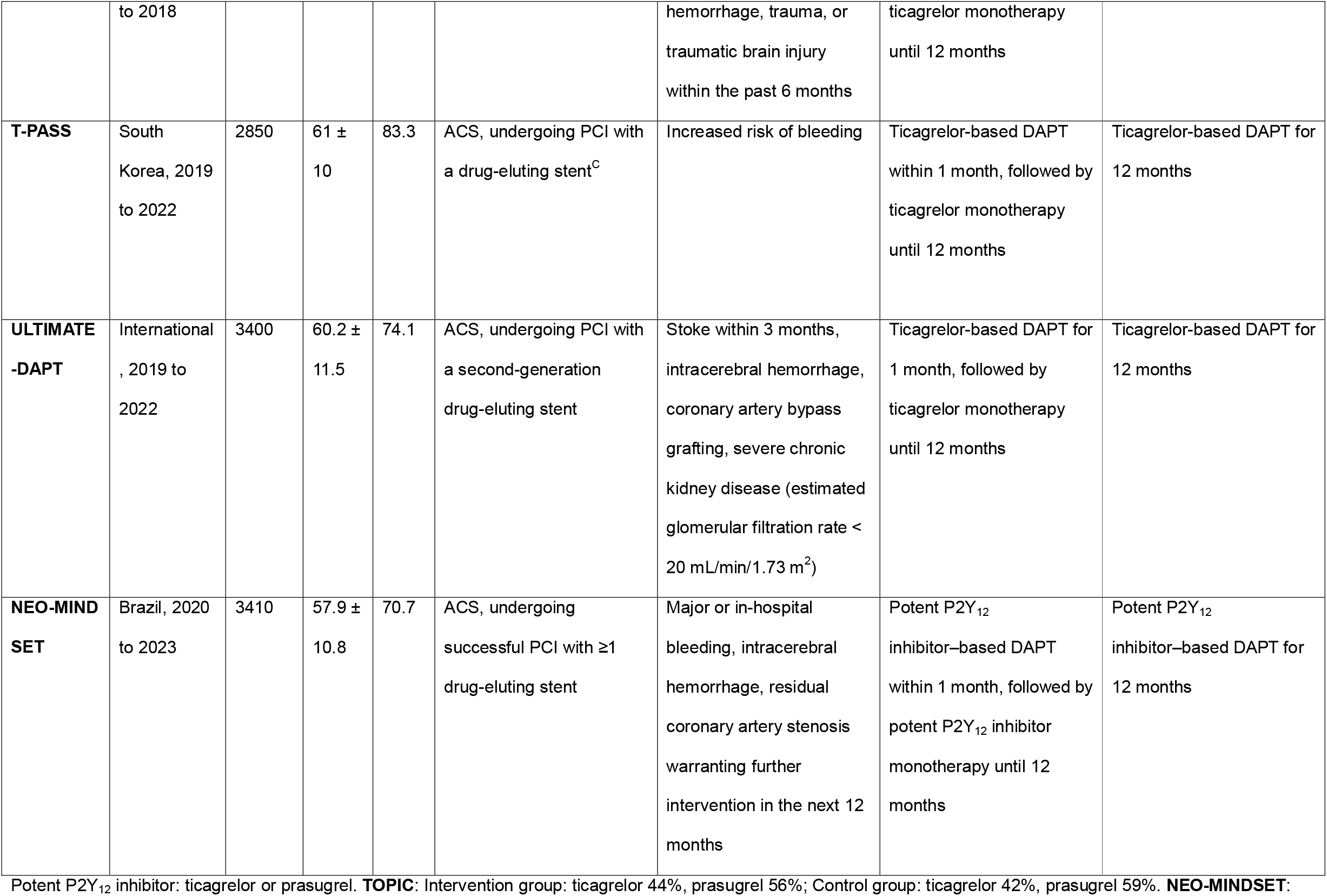
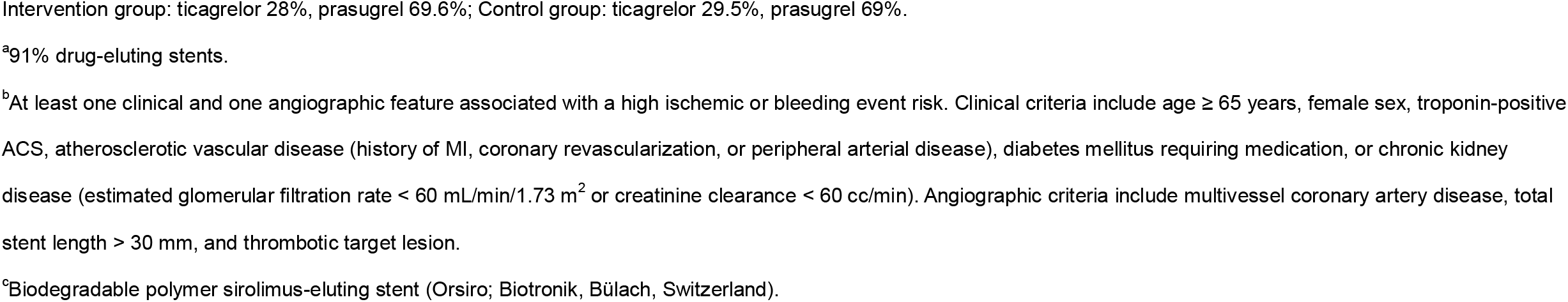
Characteristics of the Included RCTs.

Table S3 presents a summary of our risk-of-bias assessment results. Overall, the included RCTs demonstrated high methodological quality. Of the 7 included RCTs, 2 were judged as having a low risk of bias, whereas 5 were judged as having some concerns. The main reason for this judgment of “some concerns” in these 5 RCTs was their open-label design. Although this design is unlikely to affect robust clinical outcomes (eg, all-cause mortality or MI), it may introduce bias into clinician-driven endpoints, such as revascularization. However, because revascularization was not quantitatively analyzed in this network meta-analysis, the overall impact of this limitation on our pooled results is likely minimal.

### Efficacy Endpoints

In terms of the primary efficacy endpoint, MACE risk did not significantly differ between the 3 strategies in pairwise comparisons through a frequentist analysis: direct comparison of the de-escalation strategy with the standard strategy (RR = 0.71, 95% CI = 0.4-1.25, *P* =.232), direct comparison of the abbreviation strategy with the standard strategy (RR = 0.97, 95% CI = 0.8-1.19, *P* =.78), and indirect comparison of the de-escalation strategy with the abbreviation strategy (RR = 0.73, 95% CI = 0.4-1.33, *P* =.302). Moderate heterogeneity was observed across the included studies for this outcome (*τ*^2^ = 0.02, I^2^ = 40%, *P* =.15). Bayesian analysis revealed similar results for the comparison of the de-escalation strategy with the standard strategy (RR = 0.71, 95% CrI = 0.37-1.31), the comparison of the abbreviation strategy with the standard strategy (RR = 0.97, 95% CrI = 0.75-1.21), and the indirect comparison of the de-escalation strategy with the abbreviation strategy (RR = 0.73, 95% CrI = 0.37-1.44).

Secondary efficacy endpoints, including all-cause mortality, cardiovascular mortality, MI, stent thrombosis, and stroke, did not significantly differ between the 3 strategies in pairwise comparisons. These results were consistent across frequentist and Bayesian analyses (Table 3).

**Table 3.**
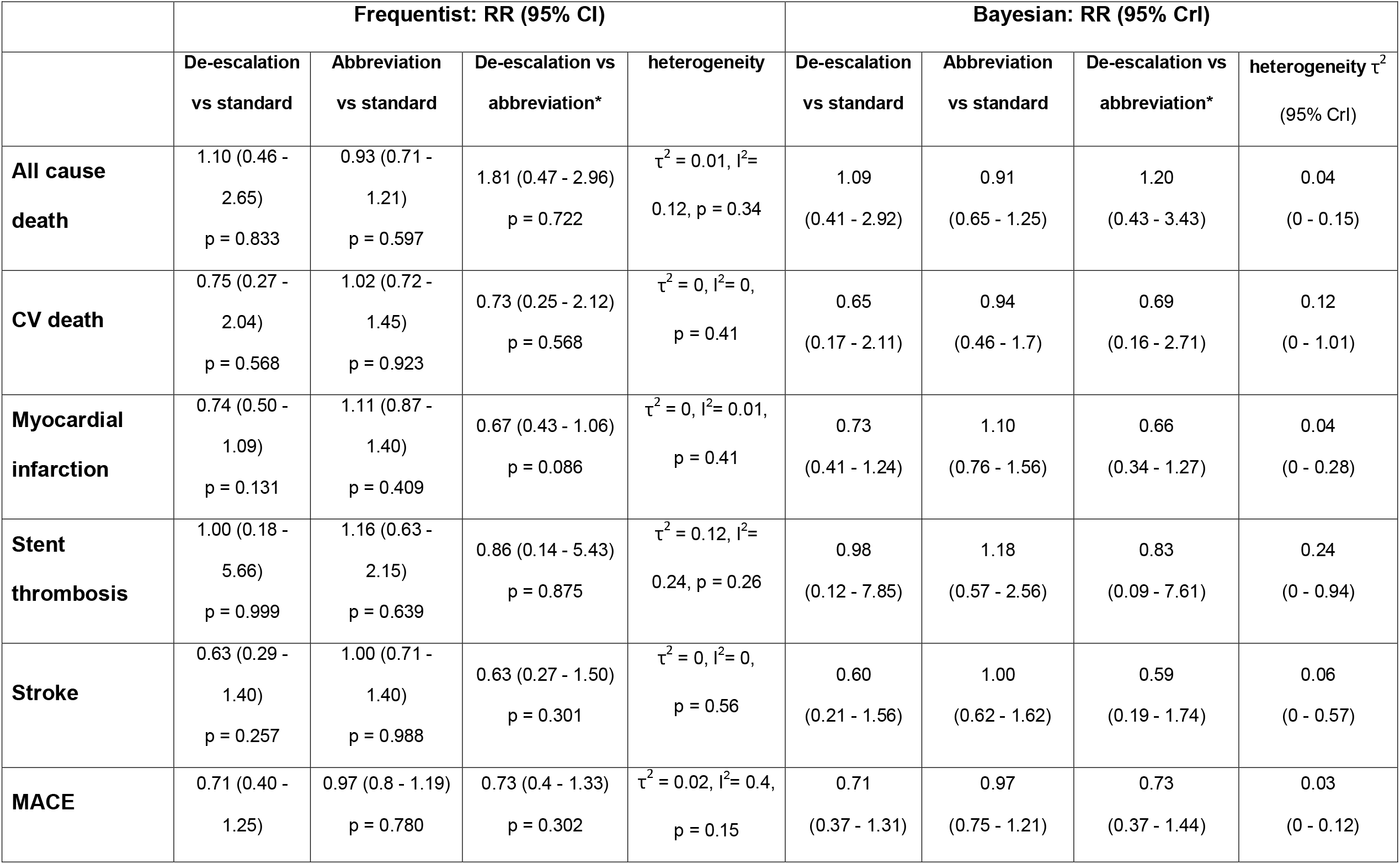
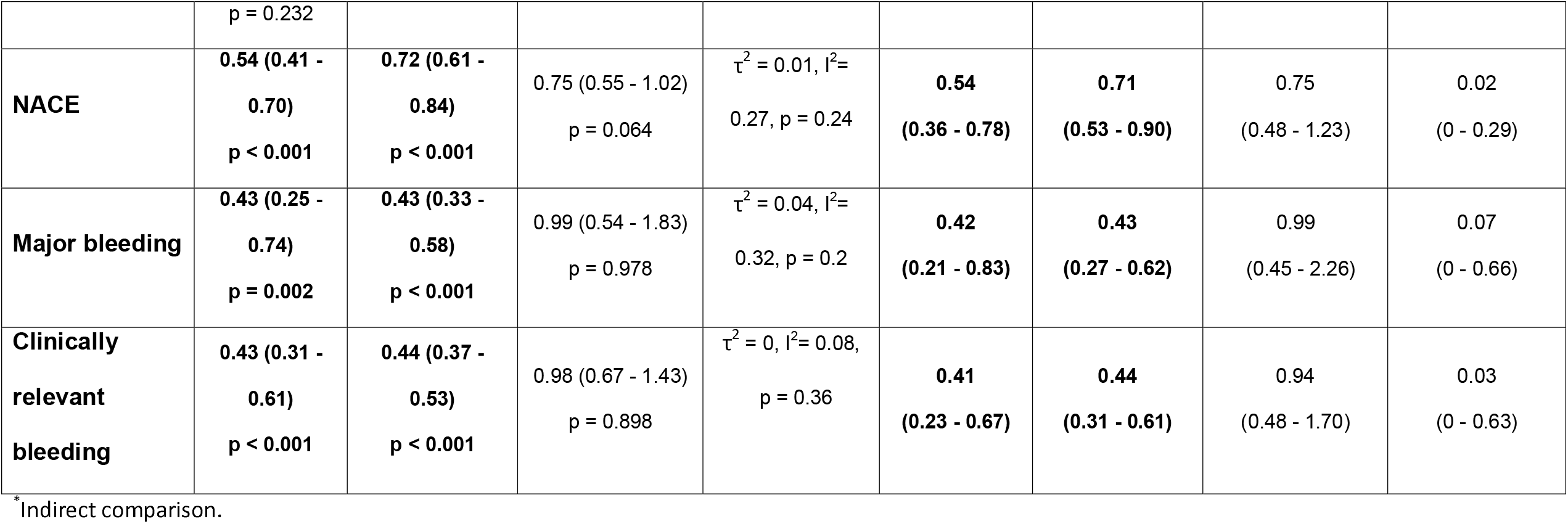
Pairwise Comparison of the 3 Strategies Through Frequentist and Bayesian Analyses.

### Safety Endpoints

In terms of the primary safety endpoint, the risk of major bleeding significantly differed in direct comparison of the de-escalation strategy with the standard strategy (RR = 0.43, 95% CI = 0.25-0.74, *P* =.002) and of the abbreviation strategy with the standard strategy (RR = 0.43, 95% CI = 0.33-0.58, *P* <.001) in frequentist analysis. However, it did not significantly differ in indirect comparison of the de-escalation strategy with the abbreviation strategy (RR = 0.99, 95% CI = 0.54-1.83, *P* =.978). Moderate heterogeneity was observed across the included studies for this outcome (*τ*^2^ = 0.04, I^2^ = 32%, *P* =.2). Similar results were observed in Bayesian analysis for this outcome: direct comparison of the de-escalation strategy with the standard strategy (RR = 0.42, 95% CrI = 0.21-0.83), direct comparison of the abbreviation strategy with the standard strategy (RR = 0.43, 95% CrI = 0.27-0.62), and indirect comparison of the de-escalation strategy with the abbreviation strategy (RR = 0.99, 95% CrI = 0.45-2.26).

In terms of the secondary safety endpoint, the risk of clinically relevant bleeding also significantly differed in pairwise comparisons of the de-escalation strategy with the standard strategy (RR = 0.43, 95% CI = 0.31-0.61, *P* <.001) and the abbreviation strategy with the standard strategy (RR = 0.44, 95% CI = 0.37-0.53, *P* <.001) in the frequentist analysis. However, it did not significantly differ in the indirect comparison of the de-escalation strategy with the abbreviation strategy (RR = 0.98, 95% CI = 0.67-1.43, *P* =.898). Low heterogeneity was observed for this outcome across the included studies (τ^2^ = 0, I^2^ = 8%, *P* =.36). Similar results were observed in Bayesian analysis: direct comparison of the de-escalation strategy with the standard strategy (RR = 0.41, 95% CrI = 0.23-0.67), direct comparison of the abbreviation strategy with the standard strategy (RR = 0.44, 95% CrI = 0.31-0.61), and indirect comparison of the de-escalation strategy with the abbreviation strategy (RR = 0.94, 95% CrI = 0.48-1.70).

### NACEs

The risk of NACEs significantly differed in direct comparisons of the de-escalation strategy with the standard strategy (RR = 0.54, 95% CI = 0.41-0.70, *P* <.001) and the abbreviation strategy with the standard strategy (RR = 0.72, 95% CI = 0.61-0.84, *P* <.001) in frequentist analysis. However, it did not significantly differ in the indirect comparison of the de-escalation strategy with the abbreviation strategy (RR = 0.75, 95% CI = 0.55-1.02, *P* =.064). Moderate heterogeneity was observed across the included studies for this outcome (τ^2^ = 0.01, I^2^ = 27%, *P* =.24). Bayesian analysis revealed similar results: direct comparison of the de-escalation strategy with the standard strategy (RR = 0.54, 95% CrI = 0.36-0.78), direct comparison of the abbreviation strategy with the standard strategy (RR = 0.71, 95% CrI = 0.53-0.90), and indirect comparison of the de-escalation strategy with the abbreviation strategy (RR = 0.75, 95% CrI = 0.48-1.23).

### Rankograms of Treatment Strategies

Figures 2 and 3 illustrate the rankograms of the treatment strategies used in the frequentist and Bayesian analyses, respectively. In terms of the primary efficacy endpoint and net clinical benefit, the de-escalation strategy consistently ranked highest, followed by the abbreviation strategy. For major bleeding, the primary safety endpoint, the probabilities of being ranked first were comparable between the de-escalation and abbreviation strategies. Overall, these findings suggest that, among the evaluated treatment approaches, the de-escalation strategy offers a more favorable balance between efficacy and safety.

**Figure 2.**
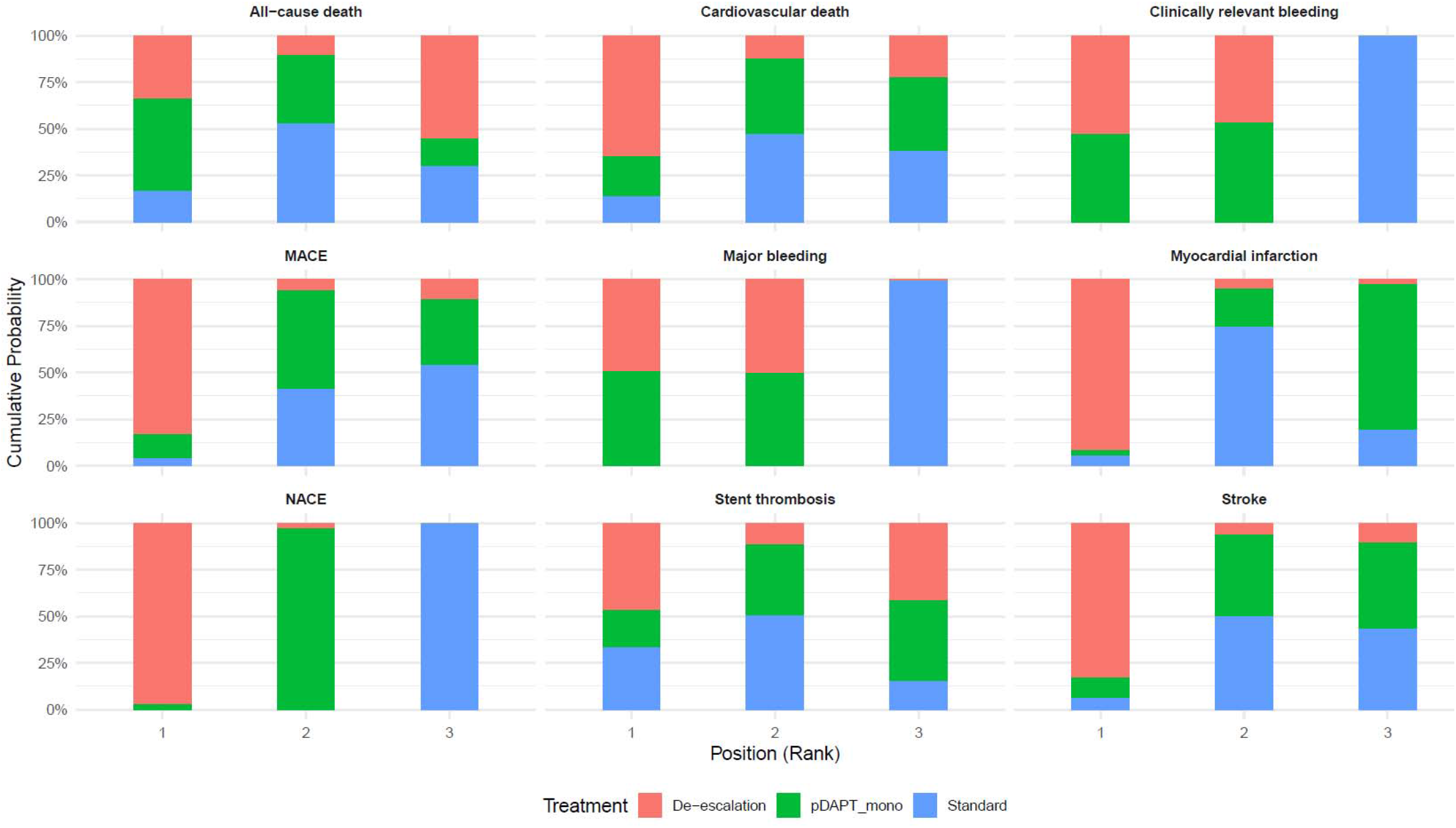
Rankograms of Treatment Strategies Based on Frequentist Analysis

**Figure 3.**
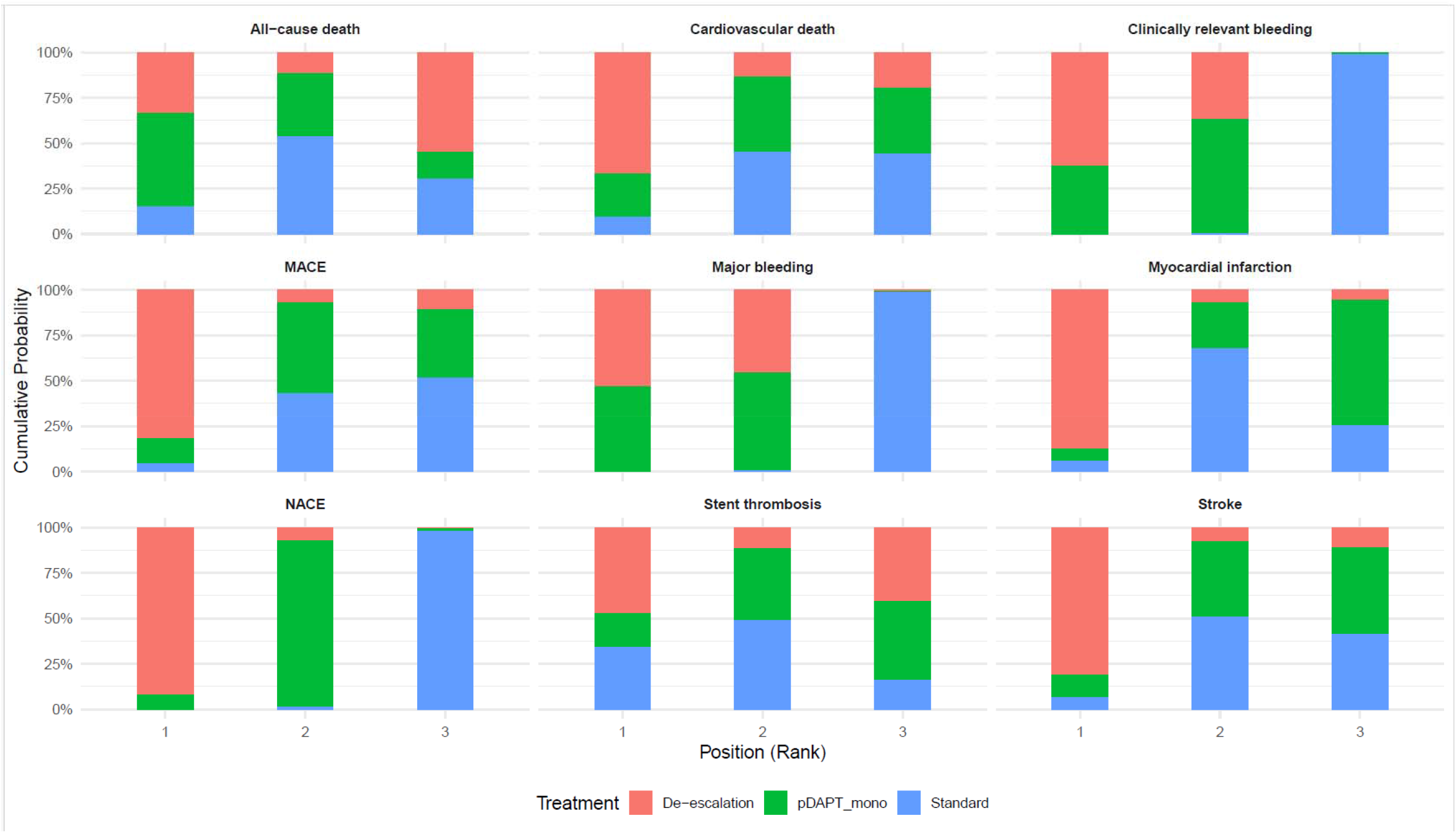
Rankograms of Treatment Strategies Based on Bayesian Analysis

### Sensitivity Analysis Results

We conducted sensitivity analyses by including only RCTs involving the administration of potent P2Y_12_ inhibitor–based DAPT within 1 or 3 months of ACS; the results are presented in Tables S4 and S5. In the sensitivity analysis limited to trials involving potent P2Y_12_ inhibitor–based DAPT within 1 month of ACS, the results of the frequentist analysis were similar to those in the overall analysis. The risk of major bleeding, clinically relevant bleeding, and NACEs significantly differed in the comparisons of the de-escalation strategy with the standard strategy and of the abbreviation strategy with the standard strategy. However, the de-escalation strategy was associated with a lower MI risk compared with the abbreviation strategy (RR = 0.54, 95% CI = 0.32-0.92, *P* =.023). In the Bayesian analysis, the posterior estimates for the abbreviation and standard strategies for major bleeding, clinically relevant bleeding, and NACEs were smaller in magnitude, with credible intervals closer to (or including) the null than in the overall analysis (Table S4).

In the sensitivity analysis limited to trials involving the administration of potent P2Y_12_ inhibitor–based DAPT for 3 months after ACS, the results of the frequentist analysis were similar to those of the overall analysis. In the Bayesian analysis, only NACEs had a 95% CrI excluding the null for comparison of the de-escalation strategy with the standard strategy (Table S5). Because the number of trials included in the sensitivity analysis was low, the degree of heterogeneity substantially increased across outcomes, as reflected by higher I^2^ and τ^2^ values and a wider τ^2^ distribution.

## Discussion

### Main Findings

The main findings of this meta-analysis are as follows: First, compared with standard potent DAPT for 12 months, short-term potent DAPT abbreviated to potent P2Y_12_ inhibitor monotherapy was significantly associated with a lower risk for major and clinically relevant bleeding and NACEs, without an increase in ischemic outcomes. Second, compared with standard potent DAPT for 12 months, short-term potent DAPT deescalated to clopidogrel-based DAPT was significantly associated with a lower risk of major and clinically relevant bleeding and NACEs, without an increase in ischemic outcomes. Third, indirect comparison of short-term potent DAPT abbreviated to potent P2Y_12_ inhibitor monotherapy and short-term potent DAPT deescalated to clopidogrel-based DAPT revealed a similar risk of MACEs, NACEs, and bleeding outcomes. In this study, we incorporated the most recent RCTs comparing clopidogrel-based DAPT and potent P2Y_12_ inhibitor monotherapy as alternative strategies after ACS. We applied narrower operational definitions for (i) deescalation to clopidogrel-based DAPT and (ii) abbreviation to potent P2Y_12_ inhibitor monotherapy after a short course of potent P2Y_12_ inhibitor–based DAPT; these definitions are stricter than those used in previous meta-analyses, and they completely align with the latest guideline recommendations. To the best of our knowledge, this is the only network meta-analysis to date that addresses the relative efficacy and safety of the 2 aforementioned strategies, providing key evidence to guide antiplatelet treatment selection for patients with ACS.

### Clinical Implications

Bleeding risk has long been a major concern in antiplatelet therapy, rendering the balance between efficacy and safety a key challenge. Both the 2025 American College of Cardiology (ACC)/American Heart Association (AHA) and 2023 European Society of Cardiology ACS guidelines endorse 12 months of potent P2Y_12_ inhibitor–based DAPT as the default post-ACS strategy (Class I, Level of Evidence A).^1,2^ In our network meta-analysis, this default approach was significantly associated with higher risks of major bleeding, clinically relevant bleeding, and NACEs, without a reduction in ischemic outcomes, compared with de-escalation to clopidogrel-based DAPT or abbreviation to potent P2Y_12_ inhibitor monotherapy after short-term potent P2Y_12_ inhibitor–based DAPT. These results are consistent with those of several recent meta-analyses, indicating that the default strategy of potent P2Y_12_ inhibitor–based DAPT for 12 months is associated with an increased bleeding risk.^13,16,17^ Taken together, these findings call into question the routine use of 12-month potent P2Y_12_ inhibitor–based DAPT as the default strategy and support the application of a more individualized, risk-adapted approach. Further adequately powered RCTs are warranted to define the types of patients who can derive net clinical benefits from prolonged potent P2Y_12_ inhibitor–based DAPT.

In this study, we applied relatively strict definitions completely aligned with the most recent guideline recommendations. This approach allowed for a more precise comparison between the strategies currently recommended in clinical practice, such as abbreviation to ticagrelor monotherapy after 1 month of potent P2Y_12_ inhibitor–based DAPT to reduce bleeding risk (Class I, Level of Evidence A) and de-escalation from potent P2Y_12_ inhibitor–based DAPT to clopidogrel-based DAPT (Class IIb, Level of Evidence B-R), in the 2025 ACC/AHA ACS guidelines.^1^ Several meta-analyses have examined the effects of post-ACS deescalation and abbreviation strategies; however, the heterogeneity in their included study populations and comparator definitions has led to uncertainty regarding differences in the efficacy and safety of potent P2Y_12_ inhibitor monotherapy and clopidogrel-based DAPT.^11,12,14–20^ Some meta-analysis classified both abbreviated DAPT or de-escalation to clopidogrel based DAPT under the same “de-escalation” arm, making it difficult to distinguish between these two approaches.^11,13,14^ Some included control arms with 12-month DAPT regimens that incorporated clopidogrel-based therapy or enrolled patient with chronic coronary syndrome, thereby limiting the interpretability of results specific to the ACS population.^12 11^There was one network meta-analysis that compared short-term DAPT or de-escalation strategy and found that de-escalation was associated with a higher major bleeding risk compared with short-term DAPT (odds ratios of 1.54 and 1.58 in 3- and 5-node networks, respectively).^19^ However, in that analysis, short-term DAPT was defined as abbreviation to either potent P2Y_12_ inhibitors, clopidogrel, or aspirin, and de-escalation was defined as reduced-dose prasugrel or ticagrelor. This methodological difference may partly explain the discrepancies among different meta-analyses.

In the present study, de-escalation to clopidogrel-based DAPT and abbreviation to potent P2Y_12_ inhibitor monotherapy were found to be associated with comparable ischemic and bleeding event risks; while rankogram analyses revealed that de-escalation to clopidogrel-based DAPT may yield the most favorable net clinical outcome. Because none of the included RCTs directly compared the 2 aforementioned strategies, we derived our findings from anchored indirect comparisons with 12-month potent P2Y_12_ inhibitor–based DAPT as the common comparator. Real-world data indicate that de-escalation or abbreviation after initial treatment with potent P2Y_12_ inhibitors is being increasingly adopted. In a large nationwide claims-based cohort study in the United States (*N* = 62423), the proportion of patients who deescalated was found to increase from 1.8% in 2010 to 12.6% in 2018, highlighting a steady secular shift toward less intensive antiplatelet strategies in clinical practice.^21^ Similarly, in a nationwide claims-based cohort study in Taiwan (*N* = 58989), 52.7% of patients initially treated with ticagrelor-based DAPT were found to deescalate to either ticagrelor monotherapy or clopidogrel-based DAPT in 9 months.^22^ Taken together, these findings underscore the practical importance of evaluating de-escalation or abbreviation strategies in contemporary ACS management.

Notably, our sensitivity analysis revealed that abbreviation to potent P2Y_12_ inhibitor monotherapy within the first month after ACS may be associated with a higher risk of MI compared with clopidogrel-based DAPT, underscoring the need to exercise caution during this high-risk period. Recent evidence suggests that the overall antiplatelet activity of potent P2Y12 inhibitor monotherapy is largely comparable with or without concomitant aspirin, except for monotherapy leading to the diminished suppression of the arachidonic acid–induced platelet aggregation pathway^23–25^ As such, whether this pathway plays a major role in the early post-ACS phase warrants further clarification.

### Reason for Not Including RCTs Involving Reduced-Dose Prasugrel as a De-escalation Strategy

Some RCTs have evaluated reduced-dose prasugrel as a de-escalation strategy.^26,27^ Consequently, several meta-analyses have grouped reduced-dose prasugrel together with clopidogrel under the de-escalation category.^18,19^ We excluded these studies because prasugrel exhibits interethnic differences in pharmacokinetics and pharmacodynamics.^28^ Compared with Western populations, East Asian populations exhibit higher plasma prasugrel concentrations and greater platelet inhibition,^29,30^ which has led to the clinical adoption of lower maintenance doses of prasugrel in these populations. Moreover, in some studies, reduced-dose prasugrel has been noted to provide stronger platelet inhibition compared with clopidogrel, ^31,32^ while East Asian trials have reported that reduced-dose prasugrel achieved comparable efficacy to ticagrelor.^33^ Therefore, we did not group reduced-dose prasugrel with clopidogrel in our de-escalation category.

### Practical Considerations

Several practical considerations related to the aforementioned strategies merit discussion. Although MACEs and NACEs remain primary factors influencing the outcomes of these strategies, economic considerations and patient adherence, which can be influenced by mild adverse effects, may also play an important crucial role.^34–36^ Potent P2Y_12_ inhibitors are relatively expensive, nevertheless, their cost-effectiveness remains an important consideration. ^35,37^ Twice-daily dosing medication may negatively affect adherence compared with once-daily medication.^38^ Additionally, clopidogrel resistance remains another key concern. Because the current study did not incorporate genotype-guided approaches, the relative efficacy of clopidogrel-based DAPT and potent P2Y_12_ inhibitor monotherapy in patients with CYP2C19 poor or intermediate metabolizers remains uncertain.

## Limitations

This study has several limitations. First, our analysis was based on trial-level rather than individual patient data. In the absence of head-to-head randomized trials, the comparisons between the de-escalation and abbreviation strategies relied on indirect evidence. Second, regional and ethnic variability in response to antiplatelet therapy across the included RCTs may limit the generalizability of our findings across different populations. Third, we did not include genotype-guided strategies in our meta-analysis; therefore, it remains unclear whether these de-escalation approaches are equally applicable to patients with CYP2C19 poor metabolizers. Finally, the included RCTs consistently excluded patients with a high bleeding risk (eg, those receiving concomitant oral anticoagulants; those with a history of intracranial hemorrhage, severe anemia, or thrombocytopenia; and those with recent major bleeding events). Thus, the antiplatelet strategy optimal for these high-risk populations remains unknown.

## Conclusion

Our findings suggest that de-escalation from potent P2Y_12_ inhibitor–based DAPT to clopidogrel-based DAPT or abbreviated to potent P2Y_12_ inhibitor monotherapy can reduce bleeding risk without compromising ischemic protection. Moreover, the risk of MACEs or NACEs does not significantly differ in anchored indirect comparisons of clopidogrel-based DAPT with potent P2Y_12_ inhibitor monotherapy. Of the evaluated strategies, clopidogrel-based DAPT may provide a favorable balance between efficacy and safety, particularly beyond the early high-risk phase after ACS. Overall, these findings underscore the potential role of individualized de-escalation strategies in optimizing post-ACS antiplatelet therapy. They also highlight the need for further relevant head-to-head RCTs and real-world data across diverse patient populations.

## Data Availability

All data used in this meta-analysis were obtained from previously published studies that are publicly available in indexed databases (PubMed, Embase and Web of Science). No individual patient data were collected for this study.

## Acknowledgements

The authors would like to thank all individuals involved in the original studies included in this meta-analysis. This manuscript was edited by Wallace Academic Editing. No additional acknowledgements are applicable.

