## Supplementary material for "Comparative Efficacy and Safety of De-escalation, Abbreviation, and Standard Potent P2Y_12_ Inhibitor–Based Dual Antiplatelet Therapy Strategies After Acute Coronary Syndrome: A Network Meta-Analysis"

**Search strategy**

### 1. Pubmed

( "acute coronary syndrome"[MeSH] OR acute coronary syndrom*[tiab] OR ACS[tiab] OR ST elevation myocardial infarction[tiab] OR non ST elevation myocardial infarction[tiab] OR STEMI[tiab] OR NSTEMI[tiab])

AND

( "Percutaneous Coronary Intervention"[MeSH] OR percutaneous coronary intervention[tiab] OR PCI[tiab] OR coronary stent*[tiab] OR drug eluting stent*[tiab] OR DES[tiab])

AND

( de-escalat*[tiab] OR deescalat*[tiab] OR "guided de-escalation"[tiab] OR step-down[tiab] OR switch*[tiab] OR abbreviat*[tiab] OR "short DAPT"[tiab] OR "shortened DAPT"[tiab] OR "early aspirin discontinuation"[tiab] OR "aspirin discontinuation"[tiab] OR aspirin-free[tiab] OR aspirin withdrawal[tiab] OR "P2Y12 monotherapy"[tiab] OR "ticagrelor monotherapy"[tiab] OR "prasugrel monotherapy"[tiab] OR "clopidogrel monotherapy"[tiab] OR "P2Y12 inhibitor discontinuation"[tiab] OR "P2Y12 receptor inhibitor discontinuation"[tiab] OR "dual antiplatelet therap*"[tiab] OR antiplatelet therap*[tiab] OR "Purinergic P2Y Receptor Antagonists"[MeSH] OR Ticagrelor[MeSH] OR ticagrelor[tiab] OR Prasugrel[MeSH] OR prasugrel[tiab] OR Clopidogrel[MeSH] OR clopidogrel[tiab] OR Aspirin[MeSH] OR aspirin[tiab] OR acetylsalicylic acid[tiab])

AND

( randomized controlled trial[pt] OR randomized[tiab] OR randomised[tiab] OR randomization[tiab] OR randomisation[tiab] OR clinical trial[pt])

AND

( "2009/01/01"[Date - Publication] : "3000"[Date - Publication])

NOT

( review[pt] OR meta-analysis[pt] OR editorial[pt] OR letter[pt] OR comment[pt] OR case reports[pt])

AND

( humans[MeSH Terms])

### 2. EMBASE:

#1. acute coronary syndrome/exp OR acute coronary syndrome:ab,ti OR ACS:ab,ti OR st elevation myocardial infarction:ab,ti OR non st elevation myocardial infarction:ab,ti OR STEMI:ab,ti OR NSTEMI:ab,ti

#2. percutaneous coronary intervention/exp OR percutaneous coronary intervention:ab,ti OR PCI:ab,ti OR coronary stent/exp OR coronary stent*:ab,ti OR drug eluting stent*:ab,ti OR DES:ab,ti

#3. (de-escalat* OR deescalat* OR "guided de-escalation" OR step-down OR switch* OR abbreviat* OR "short DAPT" OR "shortened DAPT" OR "early aspirin discontinuation" OR "aspirin discontinuation" OR aspirin-free OR aspirin withdrawal OR "P2Y12 monotherapy" OR "ticagrelor monotherapy" OR "prasugrel monotherapy" OR "clopidogrel monotherapy" OR "P2Y12 inhibitor discontinuation" OR "P2Y12 receptor inhibitor discontinuation" OR "dual antiplatelet therap*" OR antiplatelet therap*):ab,ti

#4. purinergic p2y receptor antagonist/exp OR ticagrelor/exp OR prasugrel/exp OR clopidogrel/exp OR aspirin/exp OR acetylsalicylic acid/exp OR ticagrelor:ab,ti OR prasugrel:ab,ti OR clopidogrel:ab,ti OR aspirin:ab,ti OR acetylsalicylic acid:ab,ti

#5. randomized controlled trial/exp OR clinical trial/exp OR random*:ab,ti OR randomis*:ab,ti OR randomization:ab,ti OR randomisation:ab,ti

#6. 1 AND 2 AND (3 OR 4) AND 5

### 3. Web of science

( ("acute coronary syndrome" OR ACS OR "ST elevation myocardial infarction" OR STEMI OR "non ST elevation myocardial infarction" OR NSTEMI)

AND

("percutaneous coronary intervention" OR PCI OR "coronary stent*" OR "drug eluting stent*" OR DES)

AND

(de-escalat* OR deescalat* OR "guided de-escalation" OR switch* OR step-down OR abbreviat* OR "short DAPT" OR "shortened DAPT" OR "aspirin discontinuation" OR "early aspirin discontinuation" OR aspirin-free OR "aspirin withdrawal" OR "P2Y12 monotherapy" OR "ticagrelor monotherapy" OR "prasugrel monotherapy" OR "clopidogrel monotherapy" OR "P2Y12 inhibitor discontinuation" OR "P2Y12 receptor inhibitor discontinuation")

AND

(random* OR trial) )

### 4. CENTRAL

#1 [mh "Acute Coronary Syndrome"] OR (acute coronary syndrome OR ACS OR ST elevation myocardial infarction OR STEMI OR non ST elevation myocardial infarction OR NSTEMI):ti,ab,kw

#2 [mh "Percutaneous Coronary Intervention"] OR (percutaneous coronary intervention OR PCI OR coronary stent* OR drug eluting stent* OR DES):ti,ab,kw

#3 (de-escalat* OR deescalat* OR guided de-escalation OR abbreviat* OR short DAPT OR shortened DAPT OR aspirin discontinuation OR aspirin-free OR aspirin withdrawal OR P2Y12 monotherapy OR ticagrelor monotherapy OR prasugrel monotherapy OR clopidogrel monotherapy):ti,ab,kw

#4 (TOPIC OR TALOS-AMI OR TWILIGHT OR TICO OR T-PASS OR ULTIMATE-DAPT OR NEO-MINDSET OR STOPDAPT-2 ACS OR GLOBAL LEADERS OR SMART-CHOICE):ti,ab,kw

#5 (randomized OR randomised OR "randomized controlled trial" OR RCT):ti,ab,kw

#6 #1 AND #2 AND (#3 OR #4) AND #5

Table S1. Baseline patient characteristics in each arm of the included studies

| **Study** | **Strategy** | **Number** | **Age** | **Male** | **HTN** | **DM** | **Dyslipidemia** | **STEMI** |
| --- | --- | --- | --- | --- | --- | --- | --- | --- |
| **TOPIC** | **De-escalation** | 323 | 60.6 (10.2) | 81% | 47% | 26% | 42% | 36% |
|  | **Standard** | 323 | 59.6 (10.3) | 84% | 50% | 29% | 46% | 43% |
| **TALOS-AMI** | **De-escalation** | 1349 | 60.1 (11.3) | 83.90% | 48.60% | 26.80% | 41.70% | 54.40% |
|  | **Standard** | 1348 | 59.9 (11.4) | 82.40% | 49.20% | 27.40% | 41.20% | 53.50% |
| **TWILIGHT -ACS** | **Abbreviation** | 2273 | 64.2 (10.5) | 74.50% | 67.50% | 35.60% | NA | 0% |
|  | **Standard** | 2341 | 64.2 (10.6) | 75.20% | 67.40% | 34.30% | NA | 0% |
| **TICO** | **Abbreviation** | 1527 | 61 (11) | 79% | 50% | 27% | 61% | 36% |
|  | **Standard** | 1529 | 61 (11) | 80% | 51% | 27% | 60% | 36% |
| **T-PASS** | **Abbreviation** | 1426 | 61 (10) | 84% | 47% | 30% | 74% | 40% |
|  | **Standard** | 1424 | 61 (10) | 83% | 48% | 29% | 74% | 41% |
| **ULTIMATE-DAPT** | **Abbreviation** | 1700 | 62 (11.9) | 74.40% | 62.20% | 31.80% | 69.30% | 28.70% |
|  | **Standard** | 1700 | 63 (11.1) | 73.90% | 62.50% | 31.50% | 68.10% | 27.10% |
| **NEO-MINDSET** | **Abbreviation** | 1712 | 59.5 (10.9) | 70.70% | 63.80% | 26.80% | 27.10% | 61.80% |
|  | **Standard** | 1698 | 59.8 (10.7) | 70.70% | 64.20% | 28.10% | 26.60% | 62.50% |

Table S2 definition of outcomes in each trial

| **Study** | **MACE** | **Stroke** | **NACE** | **Major bleeding** | **Clinically relevant bleeding** |
| --- | --- | --- | --- | --- | --- |
| **TOPIC** | n/a | Hemorrhagic and ischemic stroke | a composite of cardiovascular death, unplanned hospitalization leading to urgent coronary revascularization, stroke, and bleeding episodes as defined by the BARC classification >_ 2 | TIMI minor + major | BARC>=2 |
| **TALOS-AMI** | composite of CV death, MI, stroke | Hemorrhagic and ischemic stroke | Composite of cardiovascular death, mycocardial infarction, stroke, and BARC bleeding type 2, 3, or 5 | BARC >=3 | BARC 2, 3, or 5 |
| **TWILIGHT** | All-cause death, MI or stroke | Ischemic stroke | n/a | BARC >=3 | BARC 2, 3, or 5 |
| **TICO** | Major adverse cardiac  and cerebrovascular event: included the composite of death, MI, stent thrombosis, stroke, or target-vessel revascularization. | Hemorrhagic and ischemic stroke | the composite of major bleeding and major  adverse cardiac and cerebrovascular events. | TIMI minor or major  BARC 3 or 5 | n/a |
| **T-PASS** | Death, myocardial infarction, or stroke (post hoc) | Hemorrhagic and ischemic stroke | a composite of death, myocardial  infarction, stent thrombosis, stroke, and major bleeding | BARC >=3 | All bleeding (BARC type 2−5) |
| **ULTIMATE-DAPT** | Major adverse cardiovascular or cerebrovascular events | Hemorrhagic and ischemic stroke | any MACCE or any BARC bleeding [types 1, 2, 3, or 5] | BARC >=3 | BARC 2, 3, or 5 |
| **NEO-MINDSET** | Death from any cause, myocardial infarction, stroke,  or urgent target-vessel revascularization | Hemorrhagic and ischemic stroke | a composite of death from any cause, myocardial infarction, stroke, urgent target-vessel revascularization,  or a BARC type 2, 3, or 5 bleeding event. | BARC >=3 | BARC 2, 3, or 5 |

Table S3 risk-of-bias assessment

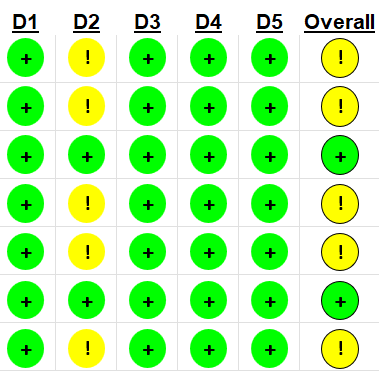

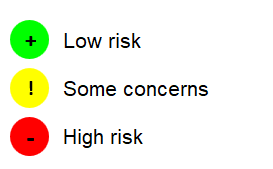

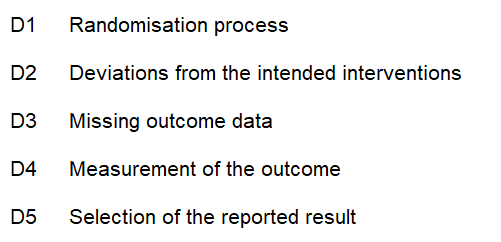

**TOPIC**

**TALOS-AMI**

**TWILIGHT**

**TICO**

**T-PASS**

**ULTIMATE-DAPT**

**NEO-MINDSET**

**Trials**

Table S4. pairwise comparison among 3 strategies:

|  | **Frequentist: RR (95% CI)** | | | | **Bayesian: RR (95% CrI)** | | | |
| --- | --- | --- | --- | --- | --- | --- | --- | --- |
|  | **De-escalation vs standard** | **Abbreviation vs standard** | **De-escalation vs abbreviation*** | **heterogeneity** | **De-escalation vs standard** | **Abbreviation vs standard** | **De-escalation vs abbreviation*** | **heterogeneity** т^2^ (95% CrI) |
| **All cause death** | 1.10 (0.46 - 2.65) p = 0.833 | 0.93 (0.71 - 1.21) p = 0.597 | 1.81 (0.47 - 2.96) p = 0.722 | т^2^= 0.01, I^2^= 0.12, p = 0.34 | 1.09  (0.41 - 2.92) | 0.91  (0.65 - 1.25) | 1.20  (0.43 - 3.43) | 0.04  (0 - 0.15) |
| **CV death** | 0.75 (0.27 - 2.04) p = 0.568 | 1.02 (0.72 - 1.45) p = 0.923 | 0.73 (0.25 - 2.12) p = 0.568 | т^2^= 0, I^2^= 0,  p = 0.41 | 0.65  (0.17 - 2.11) | 0.94  (0.46 - 1.7) | 0.69  (0.16 - 2.71) | 0.12  (0 - 1.01) |
| **Myocardial infarction** | 0.74 (0.50 - 1.09) p = 0.131 | 1.11 (0.87 - 1.40) p = 0.409 | 0.67 (0.43 - 1.06) p = 0.086 | т^2^= 0, I^2^= 0.01,  p = 0.41 | 0.73  (0.41 - 1.24) | 1.10  (0.76 - 1.56) | 0.66  (0.34 - 1.27) | 0.04  (0 - 0.28) |
| **Stent thrombosis** | 1.00 (0.18 - 5.66) p = 0.999 | 1.16 (0.63 - 2.15) p = 0.639 | 0.86 (0.14 - 5.43) p = 0.875 | т^2^= 0.12, I^2^= 0.24, p = 0.26 | 0.98  (0.12 - 7.85) | 1.18  (0.57 - 2.56) | 0.83  (0.09 - 7.61) | 0.24  (0 - 0.94) |
| **Stroke** | 0.63 (0.29 - 1.40) p = 0.257 | 1.00 (0.71 - 1.40) p = 0.988 | 0.63 (0.27 - 1.50) p = 0.301 | т^2^= 0, I^2^= 0,  p = 0.56 | 0.60  (0.21 - 1.56) | 1.00  (0.62 - 1.62) | 0.59  (0.19 - 1.74) | 0.06  (0 - 0.57) |
| **MACE** | 0.71 (0.40 - 1.25) p = 0.232 | 0.97 (0.8 - 1.19) p = 0.780 | 0.73 (0.4 - 1.33) p = 0.302 | т^2^= 0.02, I^2^= 0.4, p = 0.15 | 0.71  (0.37 - 1.31) | 0.97  (0.75 - 1.21) | 0.73  (0.37 - 1.44) | 0.03  (0 - 0.12) |
| **NACE** | **0.54 (0.41 - 0.70) p < 0.001** | **0.72 (0.61 - 0.84) p < 0.001** | 0.75 (0.55 - 1.02) p = 0.064 | т^2^= 0.01, I^2^= 0.27, p = 0.24 | **0.54**  **(0.36 - 0.78)** | **0.71**  **(0.53 - 0.90)** | 0.75  (0.48 - 1.23) | 0.02  (0 - 0.29) |
| **Major bleeding** | **0.43 (0.25 - 0.74) p = 0.002** | **0.43 (0.33 - 0.58) p < 0.001** | 0.99 (0.54 - 1.83) p = 0.978 | т^2^= 0.04, I^2^= 0.32, p = 0.2 | **0.42**  **(0.21 - 0.83)** | **0.43**  **(0.27 - 0.62)** | 0.99  (0.45 - 2.26) | 0.07  (0 - 0.66) |
| **Clinically relevant bleeding** | **0.43 (0.31 - 0.61) p < 0.001** | **0.44 (0.37 - 0.53) p < 0.001** | 0.98 (0.67 - 1.43) p = 0.898 | т^2^= 0, I^2^= 0.08,  p = 0.36 | **0.41**  **(0.23 - 0.67)** | **0.44**  **(0.31 - 0.61)** | 0.94  (0.48 - 1.70) | 0.03  (0 - 0.63) |

*indirect comparison

Table S5: Pairwise comparison among three antiplatelet strategies — Sensitivity analysis with abbreviated strategy restricted to trials with potent P2Y12 inhibitor–based DAPT maintained for 3 months after CABG

|  | **Frequentist (RR, 95% CI)** | | | | **Bayesian (RR, 95% CrI)** | | | |
| --- | --- | --- | --- | --- | --- | --- | --- | --- |
|  | **De-escalation vs standard** | **Abbreviation vs standard** | **De-escalation vs abbreviation*** | **heterogeneity** | **De-escalation vs standard** | **Abbreviation vs standard** | **De-escalation vs abbreviation*** | **heterogeneity** т^2^ (95% CrI) |
| **All cause death** | 1.10 (0.46 - 2.65) p = 0.828 | 0.68 (0.45 - 1.02) p = 0.063 | 1.62 (0.63 - 4.17) p = 0.318 | т^2^= 0, I^2^= 0,  p = 0.92 | 1.09  (0.41 - 2.91) | 0.68  (0.4 - 1.12) | 1.62  (0.54 - 4.87) | 0.03 (0 - 0.15) |
| **CV death** | 0.69 (0.21 - 2.29) p = 0.550 | 0.58 (0.17 - 2.01) p = 0.393 | 1.19 (0.21 - 6.63) p = 0.843 | т^2^= 0.17, I^2^= 0.18, p = 0.27 | 0.62  (0.14 - 2.32) | 0.57  (0.11 - 2.83) | 1.1  (0.12 - 8.96) | 0.29 (0 - 1.15) |
| **Myocardial infarction** | 0.74 (0.51 - 1.09) p = 0.129 | 0.94 (0.69 - 1.29) p = 0.716 | 0.79 (0.48 - 1.29) p = 0.338 | т^2^= 0, I^2^= 0,  p = 0.41 | 0.73  (0.39 - 1.29) | 0.89  (0.46 - 1.51) | 0.82  (0.37 - 1.96) | 0.06 (0 - 0.3) |
| **Stent thrombosis** | 1.00 (0.17 - 5.76) p = 0.999 | 0.84 (0.34 - 2.03) p = 0.693 | 1.20 (0.17 - 8.52) p = 0.859 | т^2^= 0.13, I^2^= 0.3,  p = 0.23 | 0.96  (0.16 - 5.78) | 0.81  (0.35 - 1.87) | 1.19  (0.16 - 8.57) | 0.06 (0 - 0.24) |
| **Stroke** | 0.62 (0.26 - 1.50) p = 0.290 | 1.13 (0.54 - 2.37) p = 0.742 | 0.55 (0.17 - 1.73) p = 0.307 | т^2^= 0.05, I^2^= 0.12, p = 0.32 | 0.58  (0.18 - 1.71) | 1.15  (0.44 - 3.06) | 0.5  (0.11 - 2.11) | 0.16 (0 - 0.67) |
| **MACE** | 0.71 (0.4 - 1.27) p = 0.248 | 0.85 (0.61 - 1.18) p = 0.329 | 0.84 (0.43 - 1.63) p = 0.598 | т^2^= 0.03, I^2^= 0.44, p = 0.18 | 0.71  (0.37 - 1.35) | 0.85  (0.56 - 1.24) | 0.83  (0.4 - 1.79) | 0.03 (0 - 0.13) |
| **NACE** | **0.54 (0.43 - 0.67) p < 0.001** | **0.66 (0.48 - 0.92) p = 0.012** | 0.81 (0.55 - 1.2) p = 0.294 | т^2^= 0, I^2^= 0,  p = 0.62 | **0.54**  **(0.31 - 0.91)** | 0.66  (0.31 - 1.42) | 0.81  (0.32 - 2.03) | 0.05 (0 - 0.41) |
| **Major bleeding** | **0.43 (0.23 - 0.79) p = 0.006** | **0.5 (0.31 - 0.83) p = 0.007** | 0.85 (0.39 - 1.86) p = 0.686 | т^2^= 0.08, I^2^= 0.51, p = 0.13 | 0.42  (0.17 - 1.03) | 0.50  (0.21 - 1.13) | 0.84  (0.25 – 3.00) | 0.17 (0 - 1.01) |
| **Clinically relevant bleeding** | **0.40 (0.20 - 0.77) p = 0.006** | 0.48 (0.21 - 1.10) p = 0.084 | 0.83(0.29 - 2.42) p = 0.732 | т^2^= 0.17, I^2^= 0.72, p = 0.06 | 0.39  (0.12 - 1.18) | 0.48  (0.1 - 2.26) | 0.81  (0.09 - 7.19) | 0.37 (0 - 1.53) |

*indirect comparison
